# Near point-of-care HIV viral load testing: Uptake and utilization in suburban Yangon, Myanmar

**DOI:** 10.1101/2022.07.26.22278054

**Authors:** Ni Ni Tun, Frank Smithuis, Nyan Lynn Tun, Myo Min, Myo Ma Ma Hlaing, Josefien van Olmen, Lutgarde Lynen, Tinne Gils

**Affiliations:** HIV/TB, Medical Action Myanmar, Yangon Myanmar; HIV/TB, Myanmar Oxford Clinical Research Unit, Yangon, Myanmar; Spearhead research Public Health & Primary Care, University of Antwerp, Belgium; Clinical Sciences, Institute of Tropical Medicine, Antwerp, Belgium

## Abstract

**Introduction:** HIV viral load testing in resource-limited settings is often centralized, limiting access. Near point-of-care (POC) viral load testing was introduced in Myanmar in 2017. We assessed its uptake and utilization.

**Methods:** Routine program data from three HIV clinics of Medical Action Myanmar were used. Annual viral load uptake was cross-sectionally analysed in people living with HIV (PLHIV) on antiretroviral therapy (ART) initiated between July 2009-June 2019. Attrition at two years was assessed between PLHIV with different access to viral load testing with Kaplan-Meier analysis. For those eligible for a first viral load when near POC viral load became available, a viral load cascade was constructed. We used logistic regression to explore predictors of confirmed virological failure after a first high viral load.

**Results:** Among 5271 PLHIV who started ART between July 2009-December 2019, annual viral load uptake increased significantly after near POC was introduced. Attrition in the first two years after ART initiation was not different among those eligible for a first viral load before viral load was available, after centralized laboratory-based viral load, and after near POC viral load introduction. After introduction of near POC viral load, 92% (2945/3205) of eligible PLHIV received a first viral load, a median of 2.8 years (IQR: 1.4-4.4) after initiation. The delay was 3.7 years (IQR: 2.8-5.1) and 0.9 years (IQR: 0.6-1.4) in those becoming eligible before and after near POC viral load was available, respectively. Among those with a first viral load, 95% (2796/2945) were ≤1000 copies/ml. Eighty-four % (125/149) of those with a viral load >1000 copies/ml received enhanced adherence counselling and a follow up viral load, a median of 119 days (IQR: 95-167) after the first viral load. Virological failure was confirmed in 67% (84/125), and 82% (69/84) of them were switched to second-line ART. Nine-three % (64/69) among those switched were alive on ART at end of follow-up. Having a first viral load of ≥5000 copies/ml was associated with confirmed virological failure.

**Conclusion:** Near POC viral load testing enabled rapid scale-up of viral load testing in Myanmar. PLHIV with a high viral load were adequately managed.

## Introduction

Scaling up antiretroviral therapy (ART) and access to routine HIV viral load (VL) testing are critical to control the HIV epidemic [1]. The World Health Organization (WHO) recommends VL to monitor people living with HIV (PLHIV) on ART, six months after the start of ART and annually thereafter [2]. In case of a VL above 1000 copies/mL, enhanced adherence counselling (EAC) should be performed and the VL should be repeated after three months. If the second VL is also above 1000 copies/mL despite EAC, virological failure is confirmed and prompt switch to a second-line regimen is indicated [3]. Though VL is regularly measured in developed countries, access to VL tests is still very limited in many low resource settings and this leads to delayed switching to second-line ART [4–8]. The mostly centralized VL testing demands sophisticated and expensive facilities, equipment, and skilled technicians, making it difficult and impractical to scale up. Moreover, challenges with blood collection, sample storage, and transportation often lead to delayed results [9,10].

To expand VL monitoring, WHO recommends point-of-care (POC) VL testing in priority groups since 2021 [11]. In the STREAM study, the time to the return of test results and to clinical decision making following an elevated VL reduced, and retention in care improved when POC was compared to centralized VL [12]. Near POC viral load testing, an approach using testing in laboratories close to, but not inside, treatment facilities, has shown to be feasible and to enable prompt clinical action in seven countries in sub-Saharan Africa. Compared to centralized testing, near POC also improved the turn-around time from sampling to results delivery to PLHIV and to action when high VL was identified [13,14]. For its implementation, inexpensive, low-complexity and POC assays are needed. The GeneXpert^®^ HIV-1 VL assay (Cepheid, Sunnyvale, CA; GeneXpert), is an in vitro diagnostic test which uses polymerase chain reaction (PCR) technology to quantify HIV-1 in human plasma from PLHIV [15]. Evidences have shown that the result of GeneXpert compared well with the widely used Abbott^®^ HIV-1 m2000 Real Time PCR (Abbott) [15–18]. The GeneXpert is suitable for near POC VL and was included in the WHO list of prequalified in vitro diagnostics since 2017 [19].

Myanmar is a low resource country with an overall estimated HIV prevalence of 0.6%, the second highest in Southeast Asia [20]. In 2019, Myanmar had an estimated 240 000 PLHIV, 10 000 new HIV infections and 7700 AIDS-related deaths. In the same year, 76% of PLHIV received ART, and 72% of them were virally suppressed. The epidemic is highly concentrated among key affected populations (KAP); HIV prevalence is 5.6% among female sex workers, 6.4% among men who have sex with men and 19% among people who inject drugs [20]. Routine VL monitoring was introduced in Myanmar in 2017. Until then, VL testing remained limited to centralized laboratory-based testing (Abbott Molecular; Abbott RealTi*me* HIV-1 assay) situated in two major cities, Mandalay and Yangon. Hence, access to VL testing remained limited. In 2017, only half of over 7000 newly initiated PLHIV in five regions across the country received a VL test in the first year on ART [8]. Even in six ART clinics in Yangon, a mere 58% of eligible PLHIV had received a VL test in 2019. Among the challenges encountered were sample transport, resulting in long turn-around times between sampling and reception of results [9].

GeneXpert for near POC VL testing was introduced in Yangon 2017 by the organization Medical Action Myanmar (MAM). Evidence on implementation of near POC VL testing remains limited [11]. In this study, we describe annual VL uptake and attrition according to access to VL over a 10-year period. For those eligible for VL after POC VL introduction, we present the VL cascade and predictors of having confirmed virological failure after a first high VL and EAC.

## Methods

### Study setting, design and population

The study took place at three MAM clinics in two of Yangon’s large suburban slum areas: Hlaingtharya (clinic 1 and 2) and Shwepyitha (clinic 3). These areas are generally inhabited by people with a low income, like day laborers, garment factory workers, taxi and highway drivers, and female sex workers. Many people have recently been internally displaced there from rural areas. Medical Action Myanmar (MAM) is a medical organization providing care and for PLHIV in Myanmar since 2009. MAM operates free-of-charge clinics which provide a package of services, including primary health care such as care for non-communicable diseases, reproductive healthcare, antenatal care, and tuberculosis and HIV care. Details of this HIV treatment project were described previously [21]. In short, HIV clinical care was performed by medical doctors and nurses. Counselling and adherence support through home visits were done by trained lay counsellors and outreach adherence supporters in line with the National AIDS Program and WHO HIV treatment guidelines [3,21,22]. First-line treatment consisted of a combination of two nucleoside/nucleotide reverse transcriptase inhibitors (zidovudine, tenofovir, lamivudine, abacavir) with a non-nucleoside reverse transcriptase inhibitor (efavirenz or nevirapine. Second-line regimens were composed of two nucleoside/nucleotide reverse transcriptase inhibitors, ideally which had not been used in the first-line regimen and one protease inhibitor (atazanavir/ritonavir or lopinavir/ritonavir). WHO recommended 2013 routine VL monitoring guideline was used [3]. Since 2013, one centralized Abbot HIV-1 VL testing platform was available at the National Health Laboratory (NHL) in downtown Yangon at 1 to 2 hours’ drive from the clinics. Due to the limited capacity of the NHL, only one to two samples per clinic could be sent for VL testing monthly; results could delay for over a month and many samples got lost. From January 2017, the three MAM clinics shared one near POC GeneXpert. The device was installed in a small laboratory within 30 minutes by motorbike from each clinic and was handled by trained lay personnel. Blood samples were collected at each clinic and sent daily to the laboratory. Results were provided by phone to the clinic team immediately on request, and on paper the next morning.

We used routinely collected data of PLHIV enrolled on ART in the three MAM clinics between July 2009 and June 2019. The database was closed in December 2019. The different analyses with the respective study populations are shown in fig 1.

**Fig 1.**
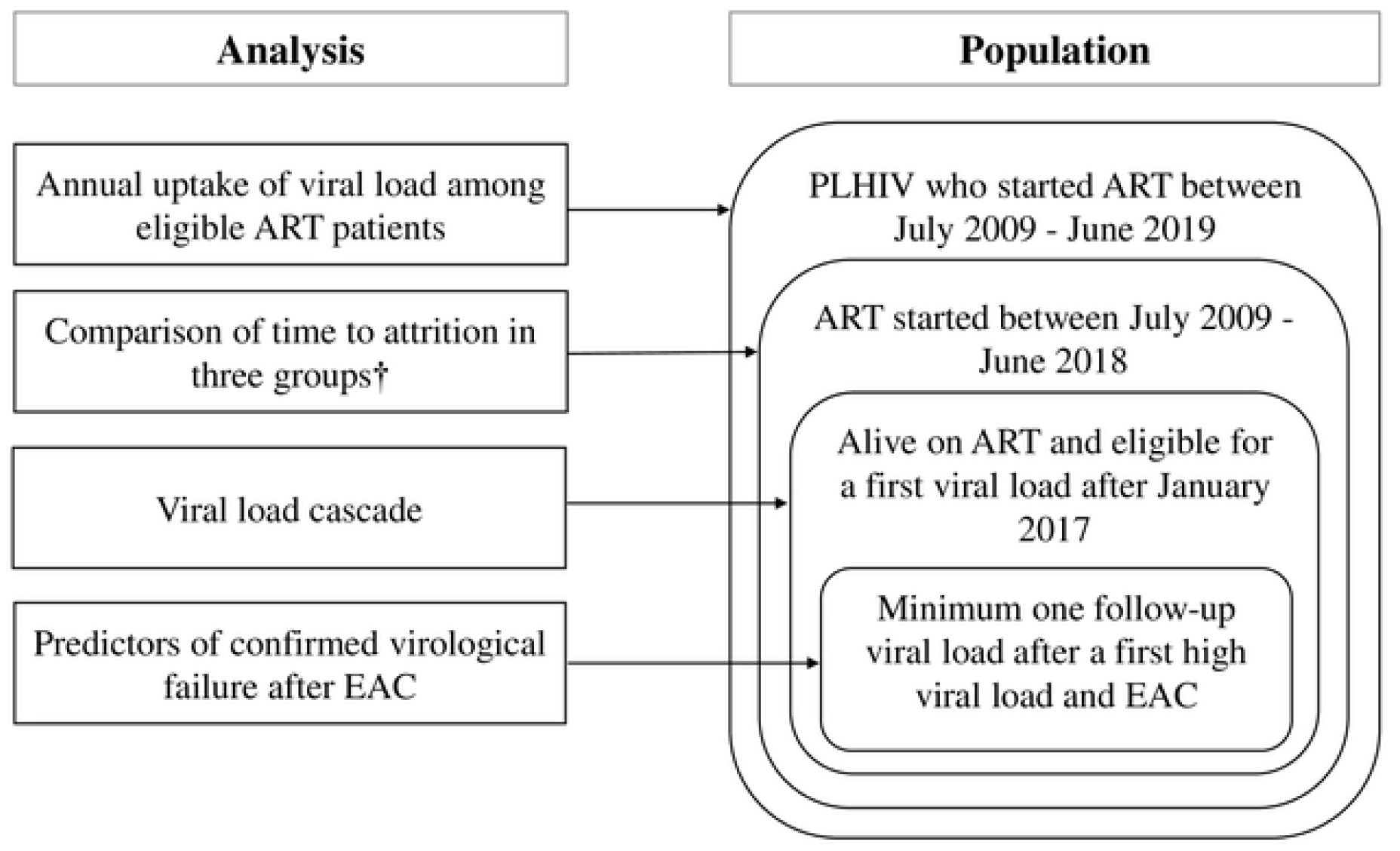
Study population per analysis performed in PLHIV on ART from three clinics in Yangon, Myanmar. ART, antiretroviral treatment; EAC, enhanced adherence counselling; PLHIV, people living with HIV/AIDS. †Group 1, six months on ART before viral load available. Group 2, eligible for viral load when laboratory-based centralized viral load available. Group 3, eligible for viral load when near point-of-care viral load available.

Annual VL uptake was analysed cross-sectionally in PLHIV eligible to have a VL done in a certain year. The remaining analyses were retrospective. Attrition during the first two years on ART was analysed among PLHIV who initiated ART between July 2009–June 2018. Three groups for comparison were defined based on the system of VL available. Group 1 consisted of PLHIV initiated on ART between July 2009–June 2012 before VL was available. Group 2 included PLHIV initiated on ART between July 2012–June 2016 who became eligible after introduction of laboratory-based centralized VL and before near POC VL. Group 3 included PLHIV initiated on ART between July 2016–June 2018, who became eligible after the introduction of near POC VL. PLHIV were considered eligible for VL when they were at least 6 months on ART, according to routine VL monitoring guideline of WHO [3]. For the VL cascade and risk factors associated with confirmed virological failure after EAC, we included those initiated ART between 1 July 2009 and 30 June 2018 (minimum potential follow-up time of 18 months on 31 December 2019) and who were eligible for a first VL from January 2017 (when POC VL was present).

### Study variables

The study used routine programme data collected from standardized patient forms in hard copy, designed for the use of FUCHIA software (Follow-Up Care of Clinical HIV infection and AIDS). Values recorded during the ART initiation visit were considered baseline for age, sex, marital status, employment, KAP status, clinic, place of origin, mode of entry, WHO stage and tuberculosis co-infection. We defined CD4 at ART initiation as the measurement taken closest to the date before ART initiation. Yearly uptake of viral load was defined as the proportion of PLHIV on ART who had at least one VL done among those who were active on ART for at least six months at the beginning of the respective year. A first VL >1000 copies/ml at least six months after ART initiation was defined as first high VL. A VL of ≤1000 copies/ml is a suppressed VL. A follow up VL after the first high VL and EAC of >1000 copies/ml is defined as confirmed virological failure. A follow up VL of ≤1000 copies/ml is a re-suppressed VL. EAC is a special form of counselling for ART patients with a first high VL in which the counsellor and patient go through the identification of barriers to adherence and strategies to overcome them. To construct the VL cascade, we assessed the proportion of ART patients who were eligible and who underwent VL monitoring, had a first high VL, had a follow up VL, had confirmed virological failure and were switched to second-line ART [3]. Outcomes were defined as follows: dead was confirmed death for any reason during the observation period. Lost to follow up (LTFU) was defined as a delay of more than three months between an expected and actual visit. The LTFU date was the last date of the clinic visit. A patient transferred to a non-MAM ART centre during the observation period was defined as transferred out (TO). Those taking ART at a MAM clinic until the end of the observation period were alive on ART. Attrition was not alive on ART due to either dead or LTFU.

### Data collection and management

Date from the laboratory database and information from the EAC counselling register were compiled with clinical data in individual patient files. These clinical files in hard copy were routinely entered into the FUCHIA database by trained data clerks. After pseudonymisation, patient-level data were extracted into Microsoft Excel. In case of discrepancies between the FUCHIA database and patient files, original records were verified, and corrections made. Data cleaning and verification was done by the first author.

### Data analysis

Categorical variables were summarized with frequencies and proportions whilst continuous variables were summarized with medians and interquartile ranges. The trend of VL uptake between was analysed with a chi-square test for trend. Kaplan-Meier techniques and a log-rank test were used to compare time to attrition of ART patients who were at least six months on ART before VL testing was introduced, after laboratory-based and before near POC, and after near POC was introduced. Follow-up time was started the day PLHIV initiated ART and ended on the date of the outcome or at database closure (31 December 2019) for those alive on ART. ART patients transferred out were censored the day they were transferred. We used stepwise multivariable logistic regression from a saturated model to explore predictors of having a confirmed virological failure after EAC. Age and sex were retained in the multivariable model, as well as variables with p-values ≤0.05. Odds ratios (OR) and 95% confidence intervals (CI) were presented. All analyses were done using Stata 14 (Stata Corp, College Station, TX).

### Ethics statement

As this is retrospective analysis, it would be impractical to collect informed consent. A consent waiver was obtained for the study by the Institutional Review Board of Institute of Tropical Medicine, Antwerp, Belgium (approval number IRB/AB/AC/058 1363/20) and Oxford Tropical Research Ethics Committee of University of Oxford.

## Results

### Annual viral load uptake

Between July 2009 and June 2019, 5271 PLHIV were enrolled on ART in three MAM clinics. VL uptake between 2010 and 2019 was 0% (0/30), 0% (0/289), 0% (0/443), 4.6% (49/1055), 5.6% (89/1579), 2.4% (49/2068), 8.2% (204/2487), 32.6% (931/2858), 91.9% (2786/3033), 94.5% (2649/2804) respectively (Fig 2). There was a significant trend towards higher uptake between 2010-2019, between 2013-2019, and between 2017-2019 (chi-square for trend p<0.001).

**Fig 2:**
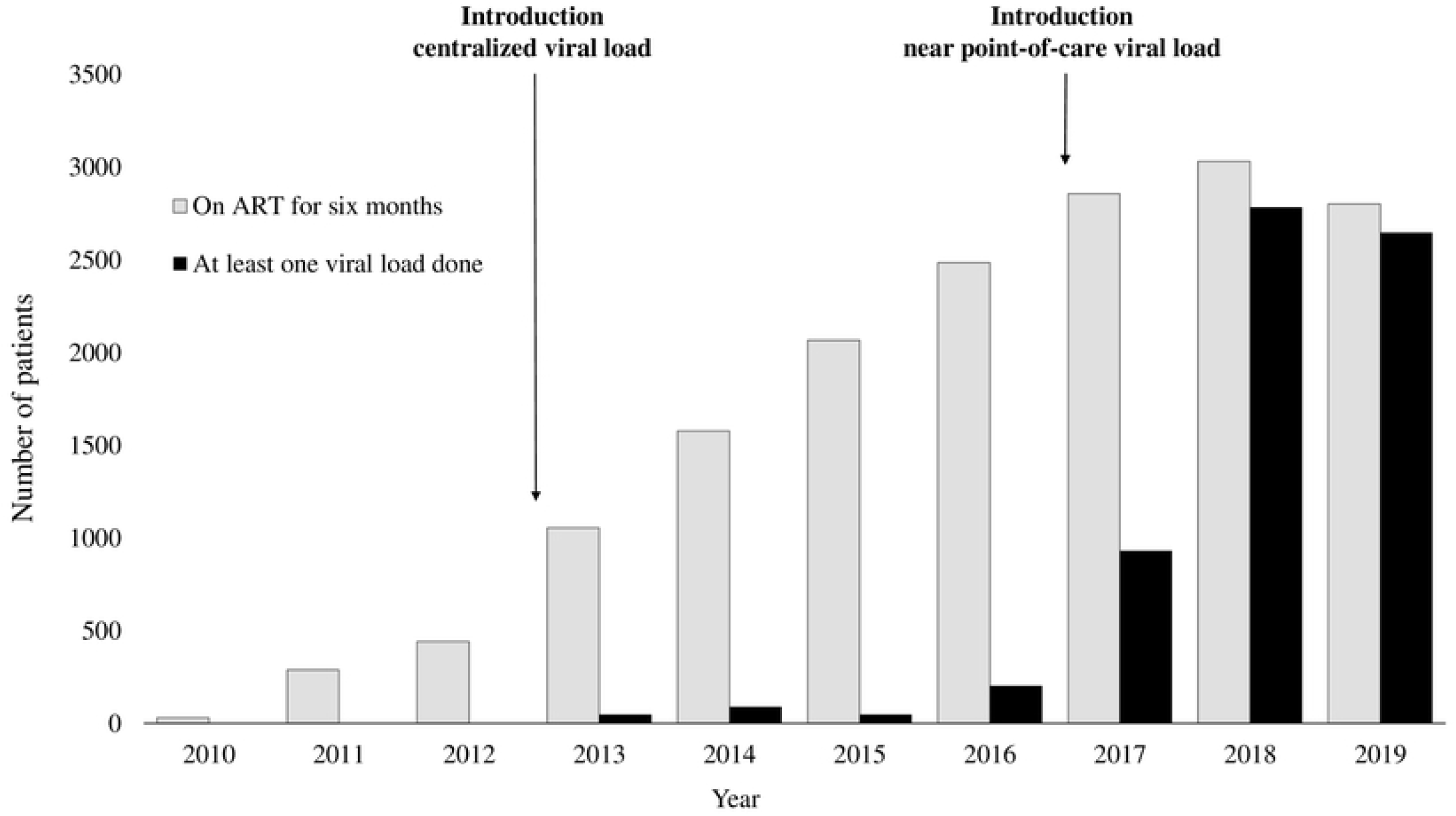
Annual viral load testing uptake among eligible PLHIV in three clinics in Yangon, Myanmar from 2010 to 2019. ART, antiretroviral treatment; PLHIV, people living with HIV/AIDS.

### Comparison of attrition

Between July 2009-June 2018, 4291 PLHIV initiated ART. Among them, 794 (18%) PLHIV were eligible to do first VL before any form of VL was introduced in Myanmar (group 1), 2386 (56%) were eligible after introduction of laboratory-based centralized VL and before the availability of near POC VL (group 2), and 1111 (26%) became eligible after introduction of near POC VL (group 3). Attrition in the first two years on ART was similar between these three groups (log-rank p-value= 0.35) (Fig 3).

**Fig 3.**
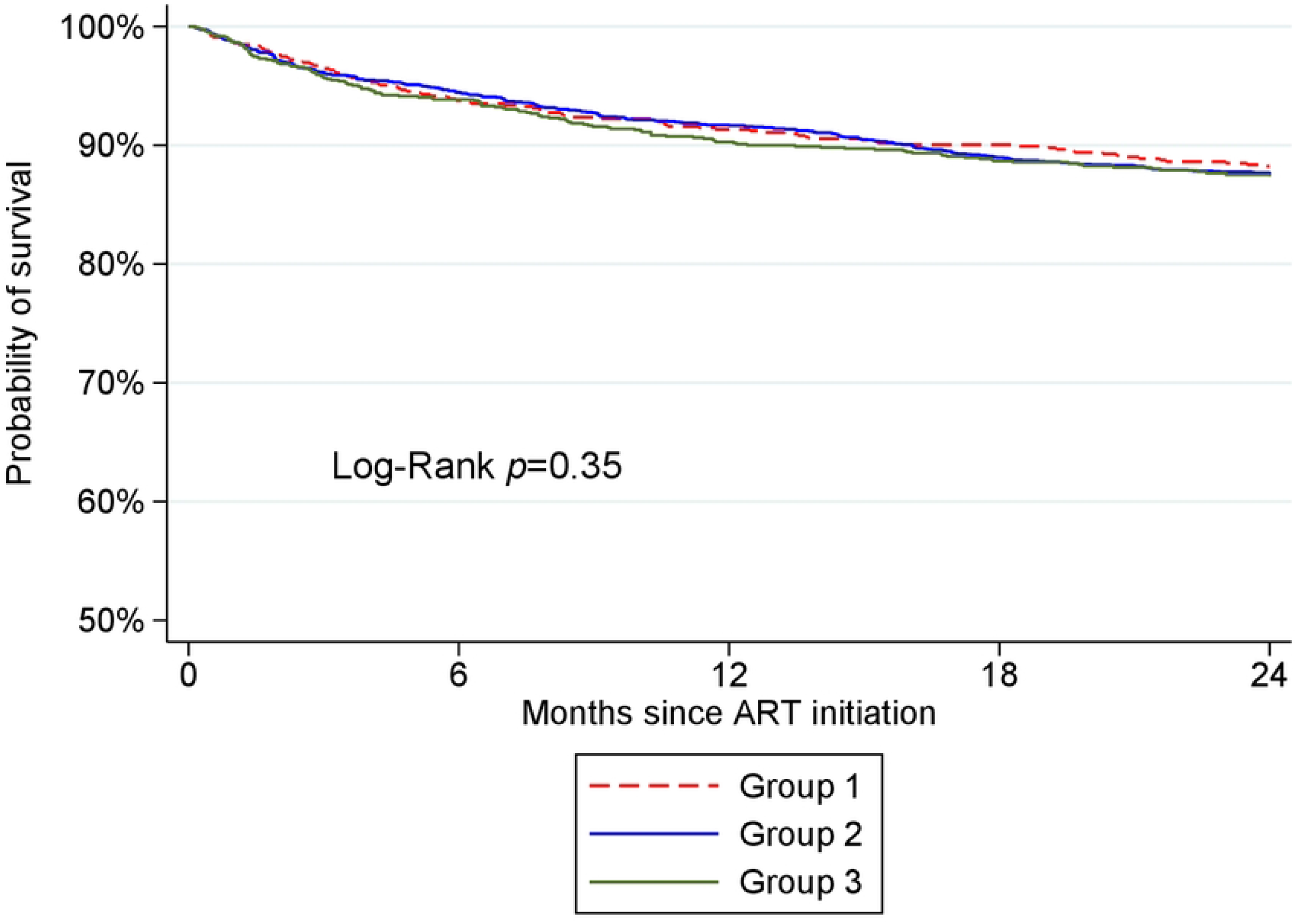
Comparing the attrition rates on ART among three patient groups in their first two years on ART. ART, antiretroviral treatment. Group 1, six months on ART when no viral load available; Group 2, eligible for viral load when centralized viral load available; Group 3, eligible for viral load when near point-of-care viral load available

### Baseline characteristics

Among 4291 patients who initiated ART between July 2009 and June 2018, 3205 were eligible for first VL testing after near POC introduction. Their characteristics at initiation are presented in table 1. The excluded 25% (1086/4291), were those who already had a first VL done (347, 32%%), those LTFU (337, 31%), were those who died (228, 21%) and were TO (174, 16%) before near POC introduction.

**Table 1:**
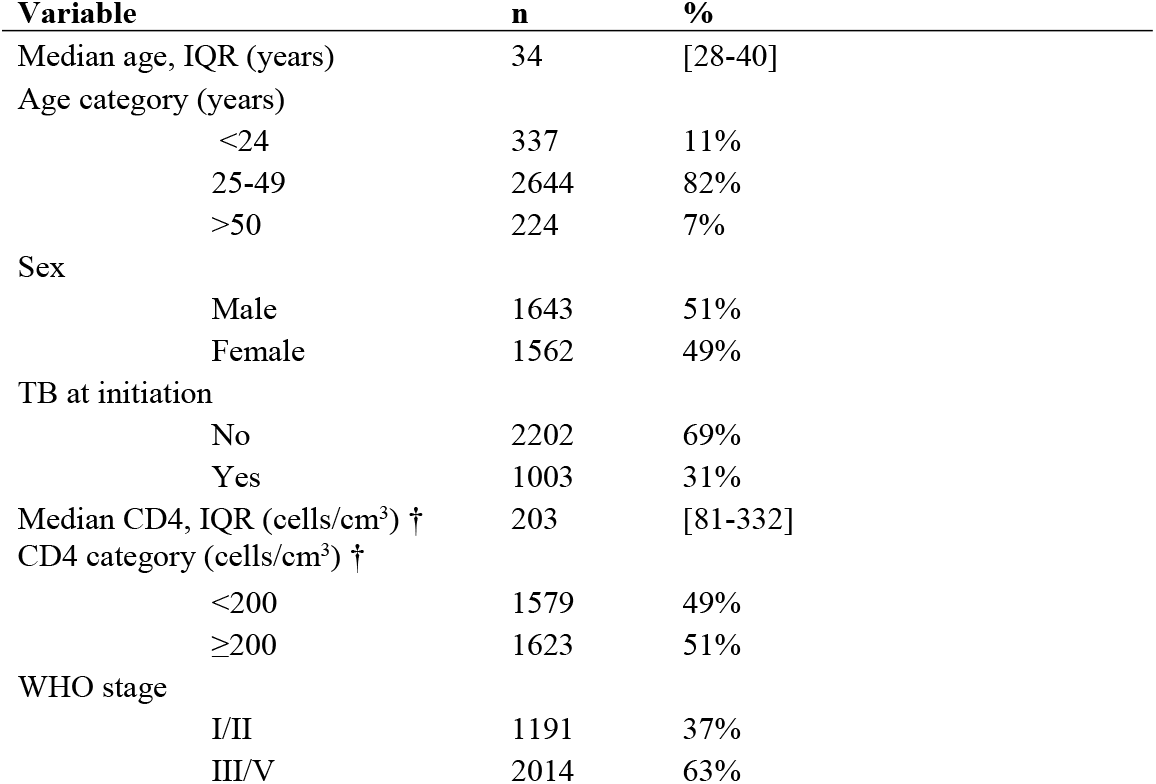
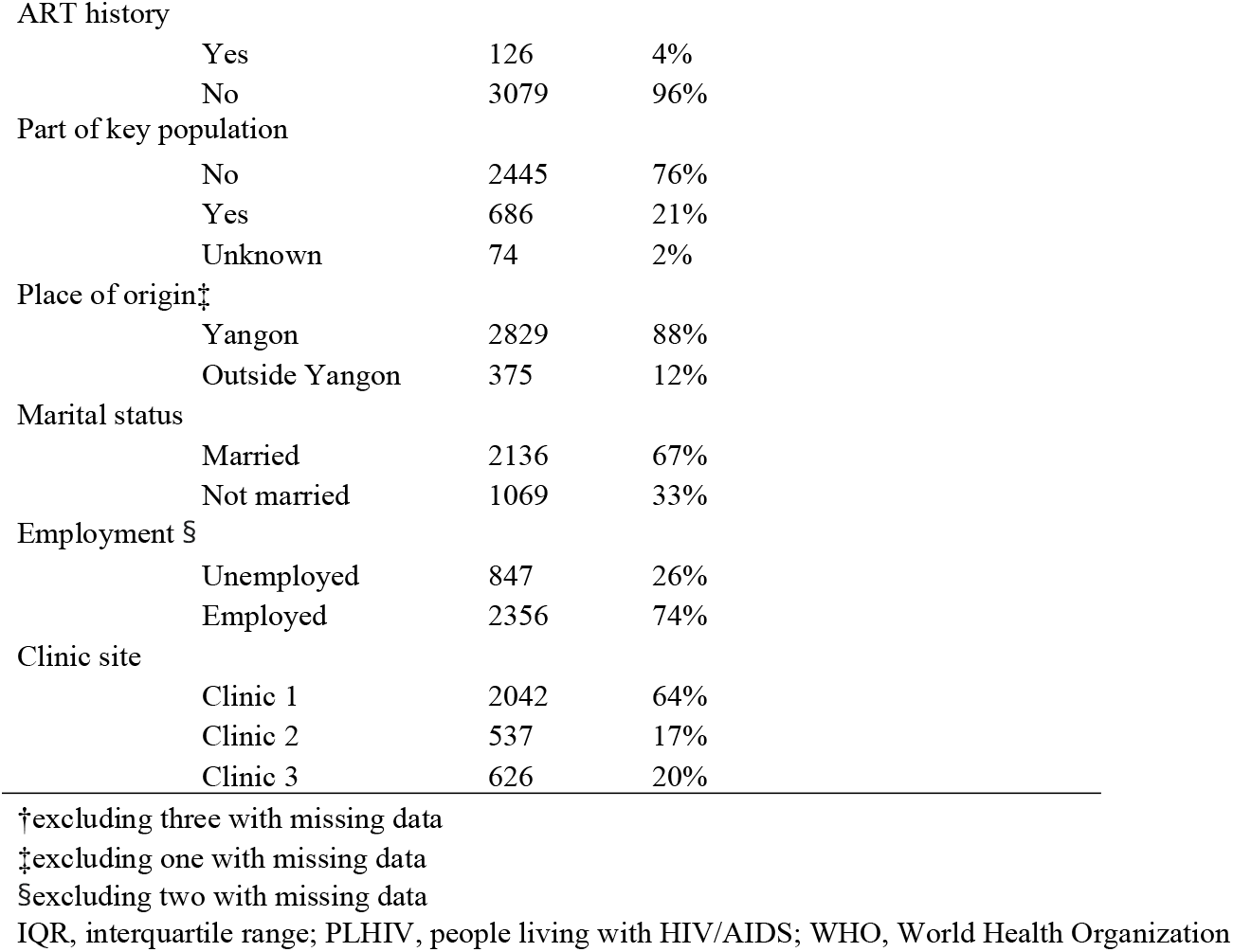
Baseline characteristics of PLHIV initiated on ART and eligible for viral load after near point-of care viral load introduction, Yangon, Myanmar (N=3205).

### HIV viral load cascade after near POC introduction

The VL cascade of those eligible for a first VL after near POC introduction is presented by Fig 4.

**Fig 4:**
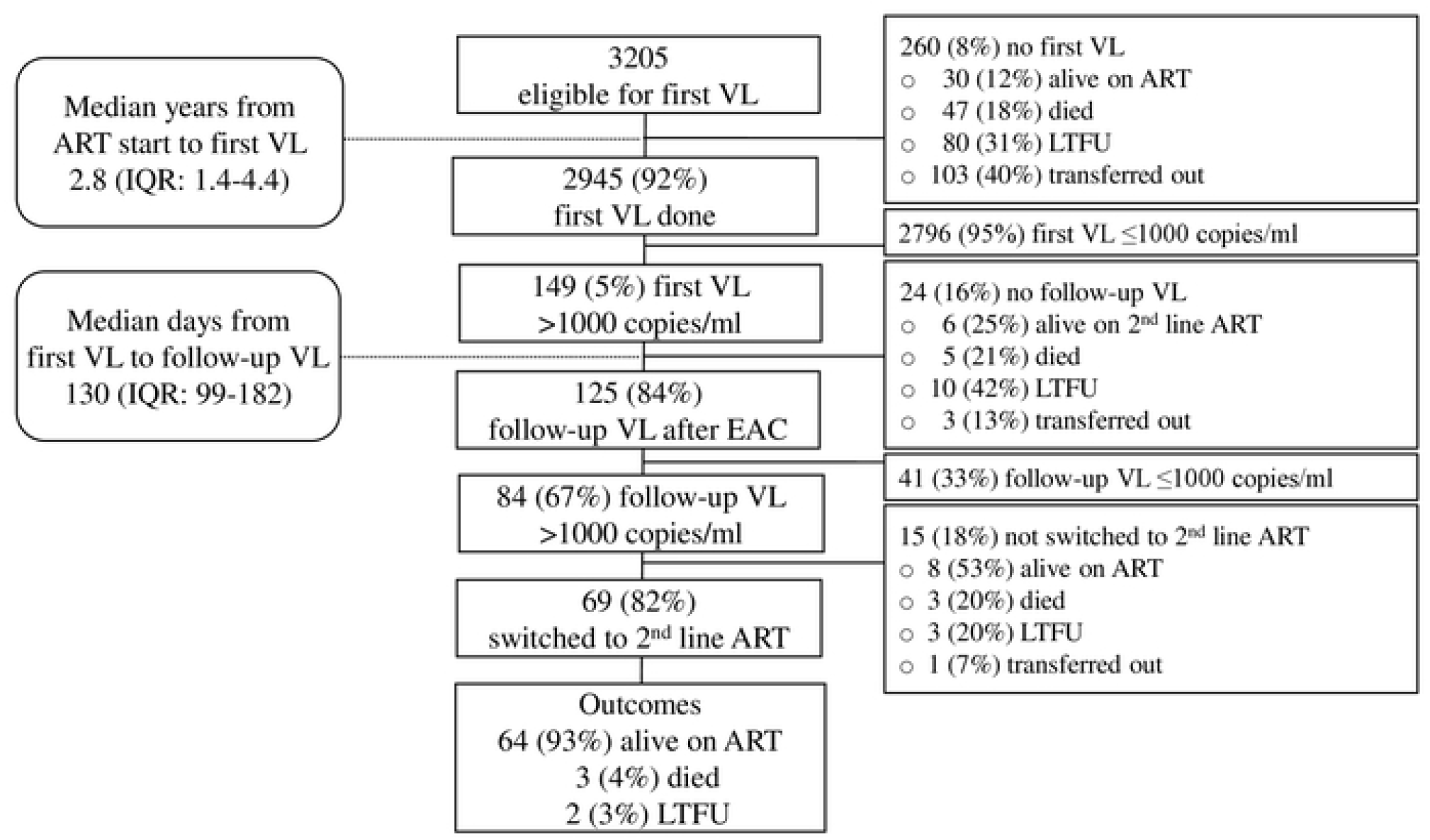
Viral load cascade of PLHIV who were enrolled on ART between July 2009 and June 2018 and eligible to do first viral load after near point-of-care viral load introduction. ART, antiretroviral treatment; EAC, enhanced adherence counselling; IQR, interquartile range; LTFU, lost to follow up; PLHIV, people living with HIV/AIDS; VL, viral load.

Of 2945 PLHIV who had done first VL test, 945 (32%) had become eligible when near POC was available (i.e. initiated ART after July 2016). For them, the median time to a first VL was 0.9 years (IQR: 0.6-1.4). For 2000 (68%) PLHIV who had started ART before July 2016, the median time to a first VL was 3.7 years (IQR: 2.8-5.1).

### Predictors of confirmed virological failure

Predictors for confirmed virological failure after enhanced adherence counselling are shown in Table 2. Among 125 PLHIV who underwent EAC and had a follow-up VL, having a first high VL result of ≥5000 copies/ml was significantly associated with confirmed virological failure in both univariate and multivariate analysis (adjusted odd ratio: 2.61 [95% CI: 1.02-6.65].

**Table 2:** Factors associated with confirmed virological failure in ART patients with enhanced adherence counselling after a first high viral load in Yangon, Myanmar.

| Variable | n | % | Univariate analysis |  |  | Multivariate analysis |  |  |
| --- | --- | --- | --- | --- | --- | --- | --- | --- |
|  |  |  | OR | 95%CI | p | aOR | 95%CI | p |
| Age |  |  | 1.00 | [0.95-1.04] | 0.86 | 1.00 | [0.96-1.05] | 0.91 |
| Sex |  |  |  |  |  |  |  |  |
| Male | 72 | 58% | ref |  |  |  |  |  |
| Female | 53 | 42% | 0.59 | [0.28-1.25] | 0.17 | 0.67 | [0.31-1.47] | 0.32 |
| Tuberculosis |  |  |  |  |  |  |  |  |
| No | 65 | 52% | ref |  |  |  |  |  |
| Yes | 60 | 48% | 2.00 | [0.93-4.31] | 0.08 |  |  |  |
| Tuberculosis at first VL |  |  |  |  |  |  |  |  |
| No | 118 | 94% | ref |  |  |  |  |  |
| Yes | 7 | 6% | 3.08 | [0.36-26.4] | 0.31 |  |  |  |
| CD4 (cells/μL) | 125 | 100% | 1.00 | [1.00-1.00] | 0.50 | | | |
| CD4 category (cells/μL) | | | | | | | | |
| <200 | 92 | 74% | ref |  |  |  |  |  |
| $\geq 200$ | 33 | 26% | 0.81 | [0.35-1.86] | 0.61 | | | |
| WHO stage |  |  |  |  |  |  |  |  |
| I/II | 27 | 22% | ref |  |  |  |  |  |
| III/V | 98 | 78% | 1.27 | [0.52-3.09] | 0.60 |  |  |  |
| ART history before baseline |  |  |  |  |  |  |  |  |
| Yes | 4 | 3% | ref |  |  |  |  |  |
| No | 121 | 97% | 6.55 | [0.66-65.0] | 0.11 |  |  |  |
| Part of key population † |  |  |  |  |  |  |  |  |
| No | 81 | 65% | ref |  |  |  |  |  |
| Yes | 42 | 34% | 0.85 | [0.39-1.87] | 0.69 |  |  |  |
| Place of origin |  |  |  |  |  |  |  |  |
| Yangon | 116 | 93% |  |  |  |  |  |  |
| Outside Yangon | 9 | 7% | 1.77 | [0.35-8.94] | 0.49 |  |  |  |
| Marital status |  |  |  |  |  |  |  |  |
| Married | 78 | 62% | ref |  |  |  |  |  |
| Not married | 47 | 38% | 1.28 | [0.59-2.74] | 0.53 |  |  |  |
| Employment |  |  |  |  |  |  |  |  |
| Unemployed | 30 | 24% | ref |  |  |  |  |  |
| Employed | 95 | 76% | 1.83 | [0.79-4.26] | 0.16 |  |  |  |
| Clinic site |  |  |  |  |  |  |  |  |
| Clinic 1 | 75 | 60% | ref |  |  |  |  |  |
| Clinic 2 | 31 | 25% | 1.54 | [0.62-3.81] | 0.35 |  |  |  |
| Clinic 3 | 19 | 15% | 3.36 | [0.90-12.56] | 0.07 |  |  |  |
| Years between ART initiation and first VL |  |  |  |  |  |  |  |  |
| <3 years | 89 | 71% | ref |  |  |  |  |  |
| ≥3 years | 36 | 29% | 0.58 | [0.26-1.29] | 0.18 |  |  |  |
| First viral load |  |  |  |  |  |  |  |  |
| <5000 copies/ml | 25 | 20% | ref |  |  |  |  |  |
| ≥5000 copies/ml | 100 | 80% | 2.79 | [1.14-6.84] | 0.03 | 2.61 | [1.02-6.65] | 0.05 |

† excluding two with missing data
Variables reported those measured at ART initiation, unless specified
aOR, adjusted odds ratio; ART, antiretroviral treatment; OR, odds ratio; viral load, VL; WHO, World Health Organization

## Discussion

This study is the first assessment of VL testing uptake and management after introduction of near POC VL in Myanmar. When near POC VL was introduced, VL uptake improved significantly, but two-year attrition was similar when comparing groups with different access to VL. Despite long delays until a first VL was done, only 5% of PLHIV had a first unsuppressed VL, and their subsequent VL cascade was adequately managed. Having a first high viral load of 5000 copies/ml or higher was associated with confirmed virological failure.

Viral load uptake increased significantly after introduction of near-POC VL as compared to laboratory-based VL. While centralized, laboratory-based VL had been introduced in 2013, VL uptake remained below 10% of those eligible until 2016. This was due in part to the fact that the two central laboratories accepted only a small number of tests and it took up to a month to get the test results. After 2017, there was a rapid scale up of VL testing, reaching 95% of those eligible in 2019. Contributing to this fast increase were the test result delivery time, which reduced from about a month to one day. This probably increased the motivation of the doctors to perform VL testing and for PLHIV have a VL test done. The relatively simple viral load procedure on GeneXpert allowed task shifting to lay workers. We were also able to secure uninterrupted supply of cartridges, which had been a challenge at introduction of centralised VL in Yangon. A reduction of results notification time was also observed after introduction of near POC VL in seven countries in sub-Saharan Africa [14].

We found no difference in attrition in the first two years after ART initiation between three groups of PLHIV with different access to VL testing (no access, limited access to centralized testing or access to near POC testing). In the STREAM randomised controlled trial, retention in care and viral suppression was significantly better in a group receiving POC VL group with tasking shifting compared to laboratory-based VL. However, retention at any clinic 12 months after ART initiation was also not found to be different between the groups [12]. In Uganda, access to viral load also did not improve survival when participants were randomized to different VL monitoring strategies [23]. We did not correct for confounders in our analysis of attrition and might thus not accurately capture the impact of near POC VL. The follow up period of two years may also be short to witness an effect of improved access to VL testing in a real-life setting. In addition, retention in care was already high before VL introduction in our cohort and remained high when laboratory-based VL was available. Measures to improve medicine adherence and retention in care should thus be continued and strengthened, regardless of the VL platform used.

The VL cascade was managed adequately after POC VL was made available. Ninety-two % of PLHIV eligible for a first VL had a VL done. This is high when compared to review data from low-and middle income countries, in which the median VL testing coverage was 74% (IQR: 46-82%) [24]. The time to a first VL was very long, but reduced when near POC became available from almost four to about one year. Despite the long waiting time for a first VL, 95% of patients who had a first VL done had a VL below1000 copies/ml. This is also high compared to other reports in resource-limited settings in Africa and Asia, including Myanmar, in which suppression rates were found between 72 and 91% [4,9,25–29]. As treatment adherence was probably good in our cohort, the long delay in VL testing did not have a large negative impact on viral load results. In earlier studies in Myanmar, high retention was also reported [21,30].

Of the PLHIV with a first high VL result, 85% received a follow up VL test after EAC. A recent review by Pham et al. reported that only 66% (median, IQR: 38-77%) of those eligible had a follow-up VL in low-and middle-income settings [24]. Only 41 (33%) of the patients had their VL re-suppressed after EAC in our study. In comparison, a systematic review from 2019 showed a higher re-suppression rate (53%) after EAC [31]. In our study, the high initial VL was probably based on true resistance and hence, patients would not benefit from EAC. Of the 84 patients whose virological failure was confirmed, 69 (82%) were switched to second line ART. This is again much higher when compared to Pham et al., reporting a switch rate of 45% (mean, IQR: 35-71) [24]. The highest switch rate came from a study from Myanmar, in which 85% of patients switched to second-line ART [8]. Like in our study, this was a cohort supported by a medical organization, in which patient follow-up might benefit from more resources compared to other settings.

We also looked at the factors associated with confirmed virological failure among the patients with a first high VL. In the 125 patients with a follow-up VL, a first high VL result of ≥5000 copies/ml was significantly associated with confirmed virological failure (adjusted odds ratio 2.61, 95%CI:1.02-6.65). Similar findings were found in Zimbabwe and Ethiopia where PLHIV with a first VL of respectively ≥5000 copies/ml and ≥10,000 copies/mL also had higher odds of virological failure after EAC. In PLHIV cohorts with high levels of adherence, high initial VL due to underlying resistance might indeed predict virological failure [29,32].

This study has several limitations. Since it is a retrospective analysis, there could be important unmeasured confounding factors. In addition, the sample size of the regression analysis was small, allowing for limited conclusions based on the results. Our clinics are managed by Medical Action Myanmar, a private medical organization receiving external funding, and results might not be generalizable to other facilities. However, all near POC sampling and analysis were managed by trained lay workers, with skills easily transferable to public health facilities. Our study also has important strengths. This study is the first one to document the impact of near POC VL on VL uptake in Myanmar, and in Southeast Asia. We present real-life results from ten years of programme implementation in a large cohort in a resource-limited setting. The principal investigator verified source files to correct incomplete data and errors that were detected in the database. In this way, despite the retrospective design, we had very little missing data.

We respond to a research gap identified by WHO addressing implementation of POC VL. The WHO conditionally recommends POC viral load, adapted to the capacity of health facilities, and while giving priority to populations at risk [11]. We provided near POC VL for all PLHIV eligible for viral load. After 2019, access to services for PLHIV deteriorated rapidly in Myanmar due to COVID-19 further due to political turmoil [33,34]. In such situations, providing services faster and closer to patients could partly mitigate impact of service disruption. Future research should show if near point-of-care viral load has this potential. In other contexts, with similar constraints in access and resources, near POC could provide an acceptable alternative to POC testing, improving time to a VL and subsequent management while limiting the burden on primary care facilities.

## Conclusion

Near POC testing rapidly increased VL uptake and lead to adequate management of ART patients with a first high VL result. In resource-constraint settings, near POC VL implementation should be considered as a priority measure to control the HIV epidemic.

## Data Availability

All relevant data are within the manuscript and its Supporting Information files.

## Competing interests

There are no conflicts of interests for all authors.

## Authors’ contributions

NNT conceived the conceived the idea and designed the study. LL, JVO & TG supported study design. FS supervised data acquisition. NNT, MM, MMMH and NLT performed data acquisition and analysis. NNT, FS, LL, JVO and TG performed data interpretation. TG supported statistical analysis. NNT drafted the first version of the manuscript, which was subsequently critically revised, edited and approved by all authors.

## Acknowledgments

We would like to thank the contribution of the study participants, and the medical and data teams from Medical Action Myanmar, and the Myanmar Oxford Clinical Research Unit, Yangon, Myanmar.

## Notes

### Competing Interest Statement

The authors have declared no competing interest.

